# Impaired alpha and beta modulation in response to social stimuli in children with autism spectrum disorder

**DOI:** 10.1101/2022.01.25.22269816

**Authors:** Lucia-Manuela Cantonas, Martin Seeber, Valentina Mancini, Aurélie Bochet, Nada Kojovic, Tonia A. Rihs, Marie Schaer

## Abstract

**Background:** Early preferential attention to biological motion is a fundamental mechanism priming the development of sophisticated skills to detect and react to social stimuli. Children diagnosed with autism spectrum disorders (ASD) demonstrate reduced visual orientation towards biological motion, however, the underlying neurobiological mechanisms are unknown.

**Methods:** We measured the neural oscillations in children with ASD (n=34, mean age 3.43 years) and age and gender matched typically developing children (TD, n=27) while watching videos of social biological (BM) and geometrical motion (GM). Their visual interest in BM stimuli was measured with eye-tracking techniques. Neural oscillations were measured as amplitude modulation of the frequency bands with the electroencephalogram and calculated as the power ratio between BM and GM conditions using scalp and brain source reconstruction analyses.

**Results:** We observed a reduced visual exploration of the BM stimuli along with (1) unchanged sensorimotor mu rhythm and (2) altered cortical alpha and beta power ratio in widespread right prefrontal areas associated with default mode and fronto-parietal networks in young children with ASD as compared to their TD peers. Furthermore, we measured significant correlations between prefrontal and posterior cingulate regions of the default mode network with the developmental quotient in both the ASD and TD groups.

**Conclusion:** We observed abnormal alpha and beta modulation of the fronto-parietal and default mode networks along with altered visual exploration of the social biological motion. These deficits represent core impairments of the disorder and may be informative in developing future behavioural and neuroregulation interventions, such as neurofeedback.

## Introduction

Typically developing infants show very early a preferential orientation of attention to biological motion. This is an early emerging fundamental mechanism priming the development of sophisticated skills to detect and interpret social stimuli [1] that facilitates the orientation towards facial expression and thus, contributes early on to social cognition development [2, 3].

Children diagnosed with autism spectrum disorders (ASD) demonstrate reduced preference for social cues from their first year of life, such as reduced attention to eyes and mouth [4, 5] and show reduced visual orientation towards biological motion [6, 7]. Behavioural studies, using eye-tracking have shown a lack of visual interest for biological motion versus scrambled motion [8], while neuroimaging studies report reduced activity in posterior superior temporal sulcus, parietal and frontal areas [9] and abnormal neural oscillatory activity over the occipito-parietal and frontal regions [10] in ASD individuals as compared to their typically developing (TD) peers during biological motion processing.

A well-studied neural measure of biological motion is the mu rhythm suppression. The mu rhythm is defined as synchronized electrical activity involving large clusters of mainly pyramidal neurons, in the brain areas responsible for voluntary movement control. It is measured non-invasively using the electroencephalogram over central sensors (C3/C4) (EEG) [11]. The mu rhythm amplitude is enhanced during rest and suppressed during motor action execution and observation, revealing the inhibition\disinhibition state of the sensorimotor cortical network [12] in alpha (8–12 Hz) as well as beta (13–30 Hz) frequency bands [13]. Several studies investigated the mu activity during action observation and imitation in children with autism and report no differences [14-17], with one study indicating that mu rhythm activity might be associated with facial imitation ability, in children with ASD and TD individuals [14]. Therefore, studies that investigate mu rhythm solely to understand the reduced visual orientation towards biological motion in children with ASD are insufficient and need complementary analyses.

In the current study, we simultaneously employed eye-tracking techniques (ET) to monitor the visual preference towards biological versus geometrical motion and high-density electroencephalogram (hdEEG) to measure the central mu rhythm suppression in a group of 34 children with ASD and 27 TD children (mean age 3.38, age range 1.7 to 5.8 years). Complementary, since the children were exposed to complex passive naturalistic biological motion stimuli with a high social content (short movies with children imitating animal-like behaviours, waving their arms, or making face grimaces), a second aim was to measure the frequency modulation of neural oscillations across all the sensors and in the source space.

## Methods and Materials

### Participants

Participants in the present study were part of a longitudinal investigation and were recruited via clinical centres specialized in ASD or French-speaking parent associations (for a comprehensive description of the cohort please refer to [18, 19]). For this study, three main inclusion criteria were applied for the ASD group: (a) a clinical diagnosis of autism spectrum disorder as assessed by an expert clinician following DSM-5 criteria [20], confirmed by scores above the cut-off at either the Autism Diagnosis Observation Schedule-Generic (ADOS-G) [21] or the Autism Diagnosis Observation Schedule, second edition (ADOS-2) [22] (the used ADOS scores represent calibrated severity scores [23]); (b) good quality EEG concomitant with a good quality ET (participants who looked at the screen for a minimum of 60% of the time), and (c) age younger than 6. The TD group was matched for age and gender with the ASD group. All TD participants were screened for neurological/psychiatric problems and learning disabilities using a medical and developmental history questionnaire and we used the ADOS-G or ADOS-2 to formally exclude the presence of any autistic symptoms. We used the same quality control threshold for the EEG and ET (criterion b above) to include them in the current study. In the final sample we included 61 participants, 34 ASD and 27 age and gender matched TD (Demographic data is presented in Table 1). More details about participant’s inclusion are provided in the Supplementary Material.

**Table 1.** Demographical and clinical data of the participants

| Characteristic | Autism spectrum disorder<br>(N34; 2F/32M) | Typically developing (N27;<br>3F/24M) | Group differences |  |  |
| --- | --- | --- | --- | --- | --- |
|  | Mean, SD | Mean, SD | t-value | df | p-value |
| Age in years | 3.43, 0.94 | 3.32, 1.31 | -0.33 | 47 | 0.73 |
| ADOS total score<sup>a</sup> | 6.8, 2 | 1.1, 0.2 | -15.84 | 32 | 1.06E-16 |
| ADOS social interaction | 6.13, 2.24 | 1.1, 0.3 | -12.50 | 33 | 2.28E-14 |
| ADOS repetitive behaviours | 8, 1.9 | 1.9, 1.8 | -11.89 | 43 | 1.72E-15 |
| Development quotient<sup>b</sup> | 89.52, 23.41 | 115.20, 16.94 | 4.84 | 56 | .00001 |

<sup>a</sup> ADOS (Autism Diagnosis Observation Schedule) scores represent calibrated severity scores [23]
<sup>b</sup> As measured with Psycho-educational Profile 3ed. or Mullen Scales of Early Learning

For each child, a developmental quotient (DQ) of children was estimated from either the Psycho-Educational Profile Third Edition (PEP-3) [24] or the Mullen Scales of Early Learning (MSEL) [25, 26]; the choice of the assessment depended on when the child was included in our longitudinal protocol (PEP-3 prior to 2015, and MSEL after). Details about the cognitive assessment are provided in the Supplementary Material.

Written informed consent was provided by all participants’ parents. The ethical committee of the University of Geneva, Switzerland, approved the study.

### Stimuli

The participants were shown two videos of dynamic biological movement and two videos of dynamic geometrical shapes, each lasting 2 minutes. There was no sound played during these 4 videos. The videos were administered in two runs and interleaved with age-appropriate cartoons [27]. For the current study, only the videos showing geometrical and biological movement were analysed. The biological motion videos included ecologically valid and complex naturalistic dynamic images of young children practising yoga, imitating animal-like behaviours, waving their arms or making face grimaces (for details [28]). For the presentations of dynamic geometric images, we used dynamic colorful geometrical shapes that resemble the classic abstract screen savers. Presentation and timing of stimuli were controlled by Tobii Studio software (Sweden, http://www.tobii.com). The visual exploration of the stimuli was measured using the TX300 Tobii eye-tracking system (sampling rate resolution of 300 Hz) and was quantified as the percentage of fixation time spent on the screen during each sequence of presented stimuli. A photocell installed on the video monitor allowed the EEG recording to be time-locked to the onset of the stimulus presentation. Children were either seating on their parents’ lap or alone in a comfortable chair while watching the videos.

### EEG acquisition and pre-processing

EEG data were continuously recorded with a sampling rate of 1000 Hz using a 129-electrode Hydrocel cap (EGI-Philips Healthcare) referenced to the vertex (Cz). Electrodes’ impedance was kept below 50 kΩ. We down-sampled the montage to a 110-channel electrode array and filtered the data between 1 and 100 Hz using Butterworth non-causal filters and a 50Hz notch filter to remove power-line contamination. Epochs with movement artefacts exceeding 100μV (per channel) were excluded. Independent Component Analysis was applied to remove eye-movement and ECG artefacts [29] using a Matlab script based on the EEGlab runica function [30] (https://sccn.ucsd.edu/eeglab/). Subsequently, noisy channels were interpolated using spherical spline interpolation [31]. The cleaned data were down sampled to 250 Hz and recalculated against the average reference. One hundred artefact-free epochs of 1 second per participant were considered as a minimum to ensure enough data quality.

Additionally, the data were subjectively evaluated between 1 to 4 according to the movement artefacts (1 being noisy data with less than 100 seconds clean epochs and therefore discarded, and 4 being very good quality data). No significant difference in data quality was measured between the two groups (t=1.08, df=58, p=0.28; ASD mean±sd, 2.8±0.4; TD mean ±sd, 3±0.3). Data pre-processing was done using Matlab (Natick, MA) and Cartool Software [32].

### EEG analysis

#### Spectral estimation

Power spectral density was computed using Welch’s method with a window size of 1 second for each electrode. Then, we computed the power ratio in decibels (dB) between conditions, i.e. biological motion observation and geometrical motion (used as baseline). Given the logarithmic nature of power ratios in dB, this ratio would be zero if the frequency-specific power in both conditions were to be the same. Negative values indicate less power or suppression in the biological motion condition, whereas a positive value indicates enhancement or greater power during the biological motion observation relative to the geometrical motion condition.

Frequency bands were defined as follows: delta, 2–4 Hz; theta, 4–8 Hz; alpha, 8-12 Hz; low beta 12–20 Hz; high beta 20-30 Hz, low gamma 30-40 Hz. For each of the six frequency bands, the averaged power ratio was calculated across all frequencies within the band for each EEG recording and each channel.

Further, we investigated the individual peaks of mu alpha (8-12 Hz) and beta (12-30 Hz) rhythmic activity. The peaks were defined in line with previous studies: the data were individually plotted topographically and the frequency at which the topography best resembled previous reports of mu activity and beta desynchronization in both children [33] and adults [34] was defined as characteristic alpha/beta frequency. The choice of selecting each participant’s mu rhythm bandwidth is important as it takes into account individual differences in the rate of maturation of the EEG signal [35].

#### Source estimation

Source estimation was applied using forward models based on realistic head geometry derived from the Montreal Neurological Institute toddler brain template of 33–44 month and conductivity data with consideration of skull thickness and cerebrospinal fluid distribution using Locally Spherical Model with Anatomical Constraints. The linear distributed inverse solution based on low resolution electromagnetic tomography was used to calculate the three-dimensional (3D) current density distribution for each solution point. The inverse solution space consisted of approximately 5000 points equally distributed in the gray matter volume [36]. The power spectral density and frequency-specific power ratios were calculated for each solution point. To reduce the dimension of the data, the grey matter was parcelled in 100 functional regions of interest using Schaefer’s parcellation model integrating local gradient and global similarity approaches using resting state functional connectivity (rs-fMRI) from 1489 participants [37]. The parcellation organizes the human cerebral cortex in 7 main networks: the sensory and motor cortices from the association cortex, the dorsal attention, ventral attention, frontoparietal control, and default networks [38]. For each subject, we extracted the power ratio for each cortical region of interest for each individual. For a detailed list of all the ROIs within the 7 networks please refer to the Supplementary Material.

### Statistical analysis

Group differences were examined using a one-way MANCOVA with the power ratio as the dependent variable and group as the independent variable, whilst adjusting for data quality and the percentage of fixation time spent on the screen during the biological and geometrical motion with Bonferroni correction for pairwise comparisons (p<.05). We first employed an exploratory analysis considering all the frequencies. The averaged power ratio across each frequency band at the scalp level (delta, 2–4 Hz; theta, 4–8 Hz; alpha, 8-12 Hz; low beta 12– 20 Hz; high beta 20-30 Hz; low gamma 30-40 Hz) for each participant was used as dependent variable. Subsequently, we considered the individual peaks of mu alpha and sensorimotor beta activity at the scalp level over central channels (C3\C4).

To evaluate the possible linear relationship between the spectral power ratio of the cortical ROIs (corresponding to the 7 main networks) and clinical and behavioural phenotypes we performed Spearman’s rank correlation coefficient with false discovery rate (FDR) correction applied to p-values using R software (Spearman-rho, two-tailed, p <.05, FDR = .05).

## Results

### Eye-tracking results

We observe no significant group effect for the amount of time spent on the moving geometrical shapes [F(1,59) = .74, p =.39, partial η2 =.01] between the two groups; however, we observe a significant group effect for the amount of time spent on the biological motion stimuli: the young children with ASD spent less time looking at the biological stimuli as compared to the TD group [F(1,59) = 15.74, p =.000, partial η2 =.21].

### EEG results

#### Individual mu rhythm

No significant group effect is measured for the individual sensorimotor alpha [C3: F(1,56) = .97, p =.32; C4: F(1,56) = .01, p =.9] and beta power ratio [C3: F(1,56) = 1.6, p =.20; C4: F(1,56) = 3.9, p =.06] over the central channels. No significant group differences are measured for the individual peak frequencies of alpha [F(1,56) = .78, p =.38)] and beta [F(1,56) = .38, p =.54] power ratio.

#### Power ratio across frequency bands

At the scalp level, the ASD group showed reduced alpha suppression over the fronto-central and right parietal sensors and enhanced high beta power ratio over the right frontal- and right parietal sensors as compared to their peers in response to biological motion relative to geometrical motion stimuli (pairwise comparisons, Bonferroni correction, p<.05). The values for each sensor showing statistical significance are displayed in the Supplementary Material. The results are displayed in Figure 2A,C. No significant differences between the groups are measured in delta, theta, low beta and low gamma frequency bands.

**Figure 1.**
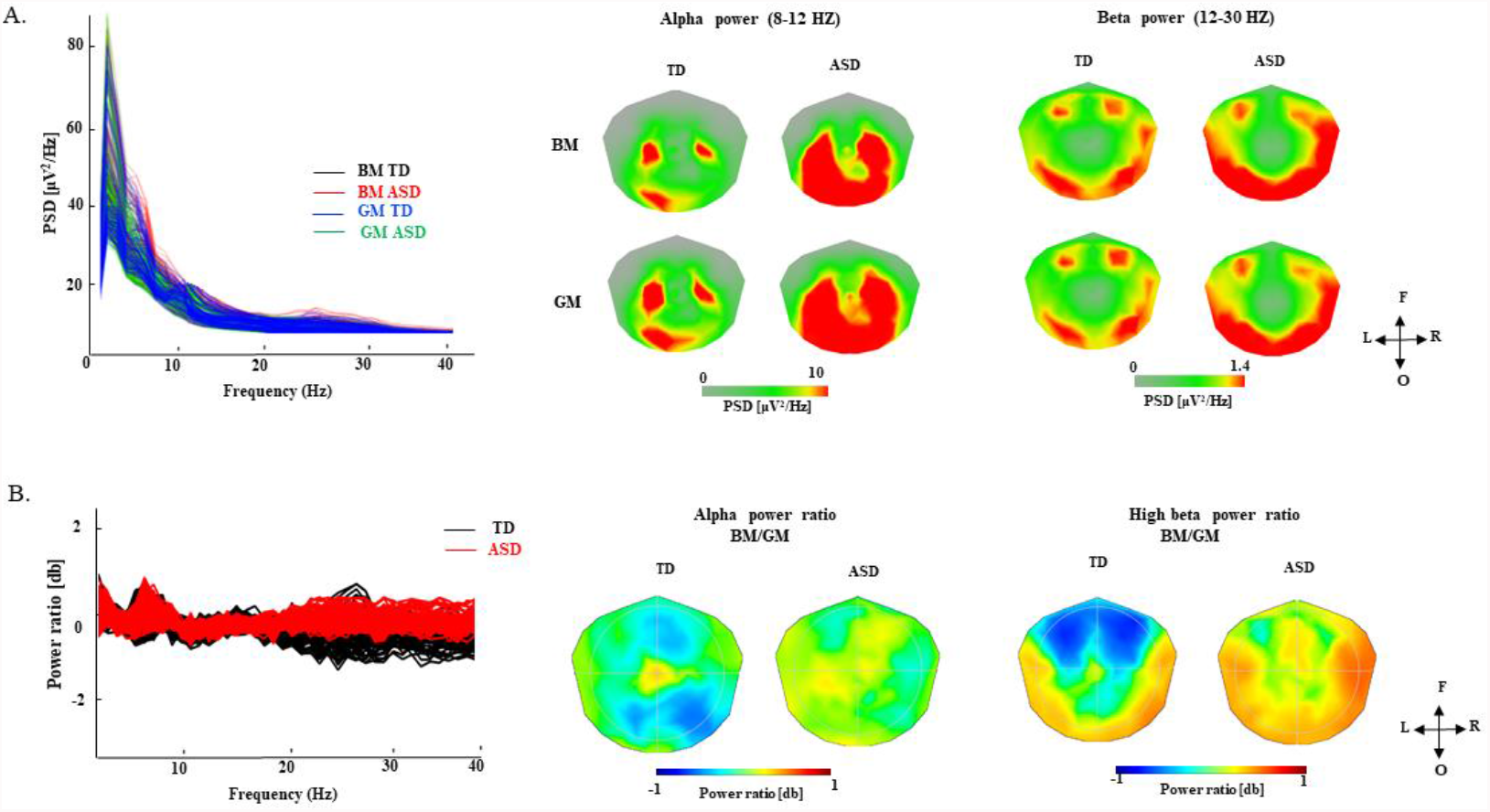
Alpha and Beta power spectrum during biological and geometrical motion. (A) Power spectrum (left) and topographic maps (right) of the mu alpha and sensorimotor beta during passive observation of biological (BM) and geometrical motion (GM) in TD and ASD groups. (B) The normalised (log10) power ratio (power of BM relative to the power of GM). On the left side, the power ratio is displayed as a butterfly plot across all frequencies (y axis) and all sensors (x axis). On the right side, the group averaged topographical representation of averaged alpha and beta ratio are displayed for TD and ASD groups.

**Figure 2.**
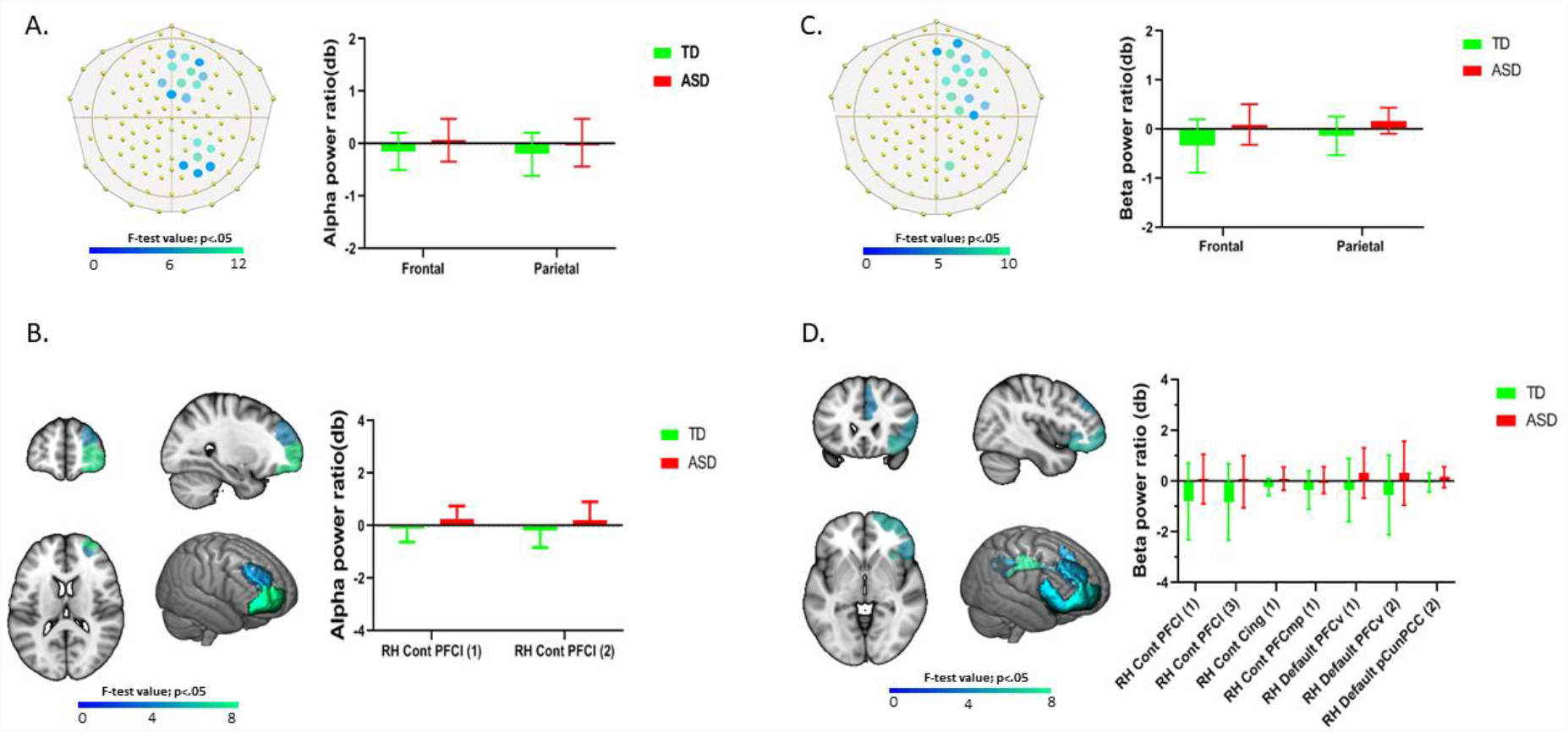
Statistical results of group differences (one-way MANCOVA, Bonferroni correction for pairwise comparisons, p<.05) for alpha and beta power ratio. Alpha power ratio: A) at the scalp level statistically significant differences over the frontal and right parietal areas (significantly different sensors are highlighted in colors according to the F-test values) and boxplot representation of the averaged significant frontal and parietal sensors; B) at the source level higher alpha power ratio in the ASD group in right lateral prefrontal areas and boxplot representation of the significant cortical areas. Beta power ratio results: C) at the scalp level statistically significant differences over right frontal and parietal areas and boxplot representation of the averaged significant sensors; D) at the source level higher beta power ratio in the ASD group in right ventral, lateral and medial prefrontal areas, praecuneus and cingulate cortex and boxplot representation of the significant cortical areas.

In the source space, the ASD group showed enhanced alpha power ratio as compared to the TD group in the right lateral prefrontal areas, corresponding to the frontoparietal control network. The children with ASD also showed reduced suppression of high beta power ratio compared to their TD peers in right lateral and medial prefrontal areas, corresponding to the frontoparietal control network, along with enhanced high beta ratio in the ventral prefrontal areas and posterior cingulate, areas that correspond to the default mode network. All the values for the ROIs showing significant statistical differences are displayed in Table 2.

**Table 2.** The values for the ROIs showing significant statistical differences.

| High Beta - ROI | Mean beta<br>power ratio<br>(sd) TD | Mean beta<br>power ratio<br>(sd) ASD | F | p | Partial eta<br>squared<br>($\eta^2$) |
| --- | --- | --- | --- | --- | --- |
| RH Control network PFCI (1) | -.77 (.25) | .06 (.22) | 5.66 | .02 | .09 |
| RH Control network PFCI (3) | -.78 (.24) | .05 (.21) | 4.53 | .04 | .07 |
| RH Control network Cingulate (1) | -.23 (.08) | .10 (.07) | 7.43 | .009 | .12 |
| RH Control network PFCmp (1) | -.34 (.13) | .04 (.11) | 4.36 | .04 | .07 |
| RH Default network PFCv (1) | -.40 (.22) | .37 (.19) | 6.02 | .02 | .09 |
| RH Default network PFCv (2) | -.56 (.28) | .33 (.24) | 4.96 | .03 | .08 |
| RH Default network PCC (1) | -.07 (.08) | .17 (.07) | 4.40 | .04 | .07 |
| Alpha- ROI | Mean alpha<br>power ratio<br>(sd) TD | Mean alpha<br>power ratio<br>(sd) ASD | F | p | Partial eta<br>squared<br>($\eta^2$) |
| RH Control network PFCI (1) | -.13 (.10) | .28 (.19) | 7.93 | .007 | .12 |
| RH Control network PFCI (2) | -.19 (.14) | .22 (.12) | 4.11 | .04 | .07 |

Abbreviations: PFCI – lateral prefrontal cortex; PFCmp – medial prefrontal cortex; PFCv – ventral prefrontal cortex; PCC- posterior cingulate cortex

The results are displayed in Figure 2 B,D.

### Correlations with clinical and developmental functioning

In the ASD group, the correlations between the alpha and beta power ratio and ADOS-2 severity scores did not survive false discovery rate (FDR) correction.

For the developmental quotient, the ASD group showed negative correlations between beta power ratio in the right medial prefrontal areas that correspond to default mode network and standard developmental scores (r =- .56, p=.001; r =- .42, p=.04; n=33), while the TD group showed positive correlations between beta power ratio in the bilateral posterior cingulate areas that correspond to default mode network and standard developmental scores (r =.50, p=.04; r = .52, p=.02; n=24). The significant correlations are displayed in Figure 3.

**Figure 3.**
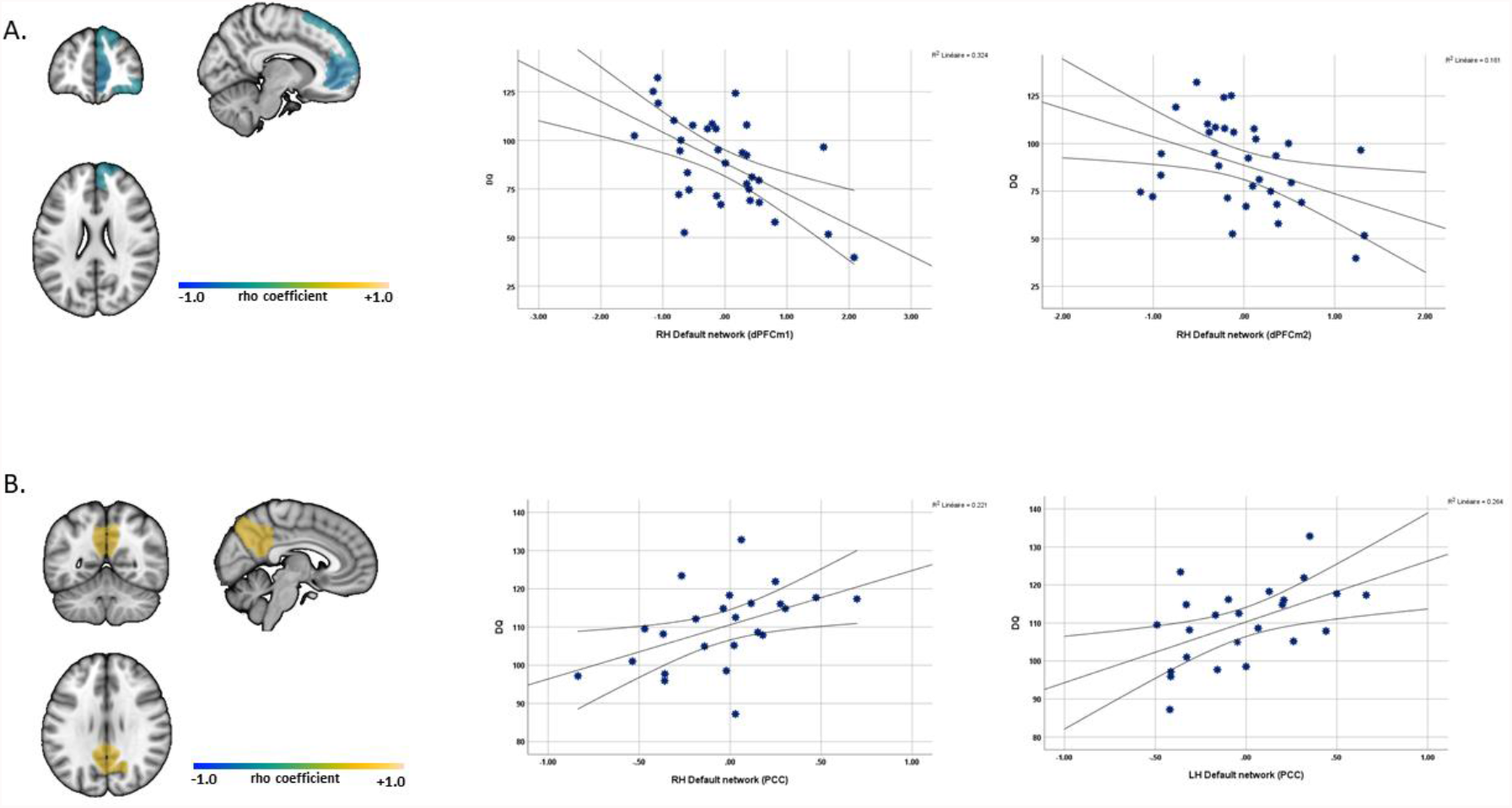
Correlation between the beta power ratio of the cortical ROIs and developmental quotient (measured using Mullen; PEP) in the ASD group (a) and the TD group (b) (Spearman-rho, 2-tailed, p <.05, false discovery rate correction).

## Discussion

To better understand the underlying neural correlates of reduced visual interest for biological motion in young children with ASD as compared to age-matched TD individuals, we measured the amplitude modulation in the EEG frequency bands in response to naturalistic biological movement using eye tracking and high-density electroencephalogram simultaneously.

The behavioural data showed reduced visual interest for biological motion (BM) and no difference in visual interest for geometrical motion (GM) in children with ASD as compared to TD children. These differences co-occurred with reduced alpha and beta suppression mainly over the right frontal and right parietal sensors at the scalp level and in the right prefrontal and cingulate areas, corresponding to the frontoparietal control (FPN) and default mode networks (DMN) at the source level. To our knowledge, this is the first evidence indicating alterations in the visual exploration of biological motion images (containing social stimuli) and in the alpha and beta frequency involving main regions of DM and FP networks. These results go in line with previous studies using eye-tracking techniques that showed reduced interest for social cues and naturalistic biological motion emerging early in development [5, 39, 40] and provide support to the hypothesis that social attention is altered in young individuals with ASD.

The behavioural changes co-occurred with differences in fronto-parietal alpha and beta modulation, but no significant differences in sensorimotor mu rhythm were found between the groups in response to BM relative to GM stimuli. These results suggest that children with ASD might have similar mu suppression to TD children over the sensorimotor areas and support previous studies reporting unaltered mu activity in children with ASD in response to goal-directed hand movement [14, 16, 41-44] or point light display human movement [17, 45]. Thus, the mu rhythm is well suppressed during motor action observation in the ASD group revealing a potentially normal inhibition\disinhibition state of the sensorimotor cortical network.

In contrast, altered scalp alpha and beta power ratio over fronto-parietal areas are measured. Alpha modulation over the frontal regions was associated with cognitive flexibility in TD pre-schoolers (for a review [46]) and high frontal-temporal alpha synchronization was associated with visual working memory maintenance [47], showing that alpha power increased with increasing cognitive demands. Beta modulation has been mostly related to motor activity still maturing throughout the early childhood [48] and less is known about how this frequency modulates in response to dynamic biological motion or social contingencies. Soto-Icaza and colleagues (2019) measured the neural activity during self-initiating joint attention while watching color drawings of toys in children with ASD and age matched TD participants and demonstrated enhanced beta oscillatory activity in the temporoparietal region that preceded social joint attention in the TD group solely [49]. Safar et al. (2021) found decreased overall functional connectivity with age (6 to 30 years old) in the beta frequency in response to happy faces in the TD group in contrast to the ASD group [50]. These results indicate that alpha and beta oscillations play a role in social information processing and suggest that alterations of these mechanisms may underlie abnormal social brain network modulation as previously reported [28].

Our results also highlighted the involvement of two main functional networks, the frontoparietal (FPN) and default-mode (DMN) networks, when toddlers and pre-schoolers visually explored dynamic social stimuli. As compared to the TD group, the ASD group showed enhanced alpha power ratio and reduced suppression of beta power ratio in right lateral and medial prefrontal areas, corresponding to the FPN, along with enhanced beta ratio in the ventral prefrontal and posterior cingulate areas corresponding to the DMN. These results indicate that children with ASD deploy higher alpha and beta activity in response to BM relative to GM stimuli, underlying higher cognitive demands during social information processing as compared to their peers. The main functions of these networks are poorly understood in early development and undergo significant remodelling with age [51-53]. The DMN is mainly involved in self-reflective processes and social cognition [54], while the FPN is thought to be involved in reasoning ability in children [55]. In previous studies on resting state data, children with ASD exhibited increased functional connectivity within regions of DM and FP networks (for a review [56]) and also showed rightward asymmetry shifts of these networks compared with their TD peers [57]. Furthermore, Lynch et al. (2013) showed hyperconnectivity of the posterior cingulate and retrosplenial cortices with medial and anterolateral temporal cortex within the DMN that was linked with severity of social impairments in the ASD group [58]. Accordingly, the abnormal modulation of these networks may partially explain the altered visual preference for social stimuli and might be related to deficits in goal-directed seeking and valuation of social information previously described in children with ASD [4, 5, 59].

We further explored the association between alpha and beta power ratio and clinical and developmental functioning. Both groups showed associations between the DMN and the developmental quotient (DQ). The ASD group showed negative correlations between beta power ratio in the right prefrontal areas and the developmental scores, while the TD group showed positive correlations between high beta power ratio in the bilateral posterior cingulate and developmental scores. These results suggest that, in children with ASD, the prefrontal high beta enhancement is associated with lower DQ level, while in TD children high beta enhancement in the posterior cingulate is associated with better DQ level. In early development the integration (stronger within-network correlations) between the posterior cingulate and the medial prefrontal cortices of the DMN are still immature, and thus might explain the different correlational patterns between the groups. Nevertheless, the role of prefrontal and cingulate high beta power ratio within the DMN in cognitive development in early ages has not been investigated by other studies to the best of our knowledge. Only few studies investigated the link between DMN nodes and IQ in school-aged children and showed no association [60] or negative association (the precentral gyrus and the frontal pole) of the DMN regions with the IQ [61] using resting state analyses.

Finally, we would like to address an important limitation of the study. Due to lack of individual MRI data, we calculated the source estimation using forward models based on realistic head geometry derived from the Montreal Neurological Institute child brain template and the regions used in this study were all derived from adult functional imaging studies (MRI). This can conduct to lower accuracy in the source localization, and thus the current results should be interpreted with caution.

Overall, we observe a reduced visual interest in response to BM and altered alpha and beta power ratio across frontal and parietal sites on the scalp in young children with ASD as compared to their TD peers. At the source level, we also measure altered cortical alpha and beta ratio in widespread right prefrontal areas associated with fronto-parietal and default-mode networks that may be implicated in the reduced visual exploration of biological motion and lower developmental levels in the ASD group.

## Data Availability

All data produced in the present study are available upon reasonable request to the authors

## Acknowledgements

The authors would like to express their gratitude to all the families who contributed to this research and to the team members who helped acquire the data over the years.

This research was supported by a grant from the National Centre of Competence in Research (NCCR) ‘SYNAPSY-The Synaptic Bases of Mental Diseases’ financed by the Swiss National Science Foundation (SNSF, 51NF40_185897), by SNSF grants to M.S. (#163859 and #190084) and by private funding from the Fondation Pôle Autisme (http://www.pole-autisme.ch).

