## Supplementary Material for "Impaired alpha and beta modulation in response to social stimuli in children with autism spectrum disorder"

Participant’s inclusion and cognitive assessment

Participants in the present study were part of a longitudinal investigation that included mainly 3 visits for the EEG data acquisition. We were interested to analyse the EEG data acquired at the first visit of each participant. In the study were included 358 participants with a first visit, among which 247 received a confirmation of diagnostic with ADOS-2 and 74 participants were described as typically developing in the control group. Further, 154 participants did an EEG recording, and 129 participants did an EEG recording and watched all the 4 required videos containing the biological and geometric motion. In the current study only 62 participants of 129 were includes due to EEG data quality (low proportion of movement artefacts). Therefore, to increase our sample, for the participants without an EEG recording of good quality at the first visit we included their EEG recording acquired during the next two visits depending on the quality of the EEG recording (*n* = 28). Furthermore, as it is a combined eye-tracking and EEG study, and therefore to ensure that participants were sufficiently looking at videos during EEG recording, we followed standard quality check procedure and we excluded participants who looked at the screen for less than 60% of the time (*n* = 11; 4 TD, 7 ASD children). The groups were age and gender matched. For the gender since we included 3 out of 11 females in the ASD group due to data quality and the time spent on the screen, we randomly selected 3 out of 21 females (1 was later excluded due to low quality EEG data) from the TD group.

The MSEL was added later (after 2015) in the study protocol, and thus the language level of participants included initially was assessed with the PEP-3 (*n* = 18, 15ASD, 3 TD) and language level of participants included after 2015 was assessed with the MSEL (*n* = 38, 17 ASD, 21TD). The MSEL is a standardized developmental assessment used in children from the ages of 0 to 68 months. The full scale includes five subdomains: Visual Reception, Fine and Gross Motor Skills, Receptive and Expressive Language. By comparison, the PEP-3 is a standardized developmental assessment for children with developmental disorders, such as ASD, between the ages of 2 and 7.5 years old. It includes the same five domains as the MSEL and includes an additional Imitation Skills Index. Data concerning language level were missing for 5 participants.

The values for the sensors showing statistical significance for the averaged alpha and beta frequencies:

Table 3 Averaged alpha ratio

| Electrode (sensor) | Mean power ratio; TD/ASD | $F_{(1,56)}$ | p-value | Partial eta squared ($\eta^2_p$) |
|---|---|---|---|---|
| E3 | -.143/.114 | 4.16 | .460 | .069 |
| E4 | -.217/.123 | 8.19 | .006 | .117 |
| E5 | -.249/.151 | 11.39 | .001 | .169 |
| E6 | -.157/.092 | 4.25 | .045 | .070 |
| E10 | -.218/.087 | 7.13 | .010 | .113 |
| E11 | -.199/.127 | 7.06 | .010 | .112 |
| E12 | -.199/.105 | 6.18 | .016 | .099 |
| E16 | -.227/.084 | 6.58 | .013 | .105 |
| E70 | -.204/.087 | 4.30 | .043 | .071 |
| E76 | -.289/.040 | 4.28 | .042 | .071 |
| E77 | -.265/.092 | 6.98 | .010 | .111 |
| E78 | -.262/.098 | 5.03 | .029 | .083 |
| E82 | -.297/.001 | 4.58 | .037 | .076 |
| E83 | -.254/.093 | 5.29 | .025 | .086 |
| E100 | -.188/.100 | 5.89 | .018 | .099 |
| E105 | -.216/.127 | 7.02 | .010 | .111 |
| E109 | -.184/.131 | 5.64 | .021 | .092 |

Table 4 Averaged beta ratio

| Electrode (sensor) | Mean power ratio; TD/ASD | F(1,56) | p-value | Partial eta squared ($\eta^2_p$) |
|---|---|---|---|---|
| E2 | -.537/.159 | 9.61 | .003 | .147 |
| E3 | -.471/.028 | 5.15 | .027 | .084 |
| E4 | -.358/.107 | 7.19 | .010 | .114 |
| E5 | -.184/.134 | 9.33 | .003 | .143 |
| E9 | -.334/.088 | 4.67 | .033 | .077 |
| E10 | -.410/.068 | 6.28 | .015 | .101 |
| E16 | -.190/.124 | 4.40 | .040 | .073 |
| E70 | -.108/.250 | 7.32 | .009 | .116 |
| E79 | -.142/.095 | 5.87 | .019 | .094 |
| E93 | -.182/.137 | 4.56 | .037 | .075 |
| E94 | -.173/.070 | 6.34 | .015 | .102 |
| E98 | -.284/.262 | 5.36 | .024 | .087 |
| E99 | -.272/.042 | 5.26 | .026 | .086 |
| E100 | -.183/.066 | 7.28 | .009 | .115 |
| E105 | -.343/.057 | 7.71 | .007 | .121 |
| E108 | -.505/.029 | 6.31 | .015 | .101 |
| E109 | -.544/.035 | 8.44 | .005 | .131 |

The detailed list of all the ROIs within the 7 networks [1].

CC_Mid_Anterior
CC_Anterior
7Networks_LH_Vis_1
7Networks_LH_Vis_2
7Networks_LH_Vis_3
7Networks_LH_Vis_4

7Networks_LH_Vis_5
7Networks_LH_Vis_6
7Networks_LH_Vis_7
7Networks_LH_Vis_8
7Networks_LH_Vis_9
7Networks_LH_SomMot_1
7Networks_LH_SomMot_2
7Networks_LH_SomMot_3
7Networks_LH_SomMot_4
7Networks_LH_SomMot_5
7Networks_LH_SomMot_6
7Networks_LH_DorsAttn_Post_1
7Networks_LH_DorsAttn_Post_2
7Networks_LH_DorsAttn_Post_3
7Networks_LH_DorsAttn_Post_4
7Networks_LH_DorsAttn_Post_5
7Networks_LH_DorsAttn_Post_6
7Networks_LH_DorsAttn_PrCv_1
7Networks_LH_DorsAttn_FEF_1
7Networks_LH_SalVentAttn_ParOper_1
7Networks_LH_SalVentAttn_FrOperIns_1
7Networks_LH_SalVentAttn_FrOperIns_2
7Networks_LH_SalVentAttn_PFCl_1
7Networks_LH_SalVentAttn_Med_1
7Networks_LH_SalVentAttn_Med_2
7Networks_LH_SalVentAttn_Med_3
7Networks_LH_Limbic_OFC_1
7Networks_LH_Limbic_TempPole_1
7Networks_LH_Limbic_TempPole_2
7Networks_LH_Cont_Par_1
7Networks_LH_Cont_PFCl_1
7Networks_LH_Cont_pCun_1
7Networks_LH_Default_Temp_1
7Networks_LH_Default_Temp_2
7Networks_LH_Default_Par_1
7Networks_LH_Default_Par_2
7Networks_LH_Default_PFC_1
7Networks_LH_Default_PFC_2
7Networks_LH_Default_PFC_3
7Networks_LH_Default_PFC_4
7Networks_LH_Default_PFC_5
7Networks_LH_Default_PFC_6
7Networks_LH_Default_PFC_7
7Networks_LH_Default_pCunPCC_1
7Networks_LH_Default_pCunPCC_2
7Networks_RH_Vis_1
7Networks_RH_Vis_2
7Networks_RH_Vis_3

7Networks_RH_Vis_4
7Networks_RH_Vis_5
7Networks_RH_Vis_6
7Networks_RH_Vis_7
7Networks_RH_Vis_8
7Networks_RH_SomMot_1
7Networks_RH_SomMot_2
7Networks_RH_SomMot_3
7Networks_RH_SomMot_4
7Networks_RH_SomMot_5
7Networks_RH_SomMot_6
7Networks_RH_SomMot_7
7Networks_RH_SomMot_8
7Networks_RH_DorsAttn_Post_1
7Networks_RH_DorsAttn_Post_2
7Networks_RH_DorsAttn_Post_3
7Networks_RH_DorsAttn_Post_4
7Networks_RH_DorsAttn_Post_5
7Networks_RH_DorsAttn_PrCv_1
7Networks_RH_DorsAttn_FEF_1
7Networks_RH_SalVentAttn_TempOccPar_1
7Networks_RH_SalVentAttn_TempOccPar_2
7Networks_RH_SalVentAttn_FrOperIns_1
7Networks_RH_SalVentAttn_Med_1
7Networks_RH_SalVentAttn_Med_2
7Networks_RH_Limbic_OFC_1
7Networks_RH_Limbic_TempPole_1
7Networks_RH_Cont_Par_1
7Networks_RH_Cont_Par_2
7Networks_RH_Cont_PFCl_1
7Networks_RH_Cont_PFCl_2
7Networks_RH_Cont_PFCl_3
7Networks_RH_Cont_PFCl_4
7Networks_RH_Cont_Cing_1
7Networks_RH_Cont_PFCmp_1
7Networks_RH_Cont_pCun_1
7Networks_RH_Default_Par_1
7Networks_RH_Default_Temp_1
7Networks_RH_Default_Temp_2
7Networks_RH_Default_Temp_3
7Networks_RH_Default_PFCv_1
7Networks_RH_Default_PFCv_2
7Networks_RH_Default_PFCdPFCm_1
7Networks_RH_Default_PFCdPFCm_2
7Networks_RH_Default_PFCdPFCm_3
7Networks_RH_Default_pCunPCC_2

1. Yeo, B.T., et al., *The organization of the human cerebral cortex estimated by intrinsic functional connectivity.* J Neurophysiol, 2011. **106**(3): p. 1125-65.
